# Alpha-Linolenic Acid Associations with Disability and Brain Volume in Multiple Sclerosis: A Brief Replication Report

**DOI:** 10.64898/2026.01.19.26344386

**Authors:** Max Korbmacher, Kjell-Morten Myhr, Stig Wergeland, Kristin Wesnes, Øivind Torkildsen

## Abstract

**Objective:** To replicate and extend recent findings suggesting that higher serum alpha-linolenic acid (ALA) levels are associated with reduced disease activity and progression in multiple sclerosis (MS).

**Methods:** We reanalysed clinical trial data from 85 people with MS, who had serum ALA, magnetic resonance imaging (MRI), and clinical (EDSS, PASAT) assessments collected for two years, with additional follow-up at 12-years. Linear and mixed models were used to assess the relationship between ALA and clinical and MRI outcomes. Mediation analyses tested whether ALA mediated associations between brain volume or T2 lesion load, and disability.

**Results:** ALA measures were consistent over time (κ= 0.83). Higher ALA predicted lower EDSS (β = −0.41, 95% CI [−0.73, −0.08]) and larger brain volume (β = 0.22, 95% CI [0.09, 0.36]). ALA was a non-significant mediator of brain volume or lesion effects on EDSS and did not predict long-term clinical or cognitive change.

**Discussion:** We replicate prior associations between higher serum-ALA levels and reduced disability in MS and extend these by showing a beneficial association of serum-ALA with brain volume. However, ALA did not predict long-term progression, limiting its prognostic value.

## Introduction

Despite advances in multiple sclerosis (MS) treatments, many people with MS continue to experience disease progression. Emerging evidence highlights a potential role of dietary polyunsaturated fatty acids (PUFAs) in modifying MS disease activity and progression. In a recent prospective study, Cortese and colleagues^1^ reported that higher serum levels of alpha-linolenic acid (ALA), a plant-derived omega-3 PUFA, were associated with reduced risk of clinically definite MS (CDMS), fewer relapses, and slower disability progression over 11 years of follow-up. To assess the reproducibility and external validity of these results, we conducted an independent replication using long-term data from a clinical trial cohort. We followed the original study’s analysis plan but extended it to include mediation analyses for key effects.

## Method

We replicated the recent analyses by Cortese et al.^1^ by reanalysing previous clinical trial data including 85 Norwegian persons with MS.^2^ Serum alpha-linolenic acid (ALA), Expanded Disability Status Scale (EDSS) scores, Paced Auditory Serial Addition Test (PASAT), and T1 and T2-weighted MRI were sampled yearly over 2 years, and repeated at 12-year follow-up. The general data collection procedure,^2^ and more specifically, the serum sampling^3,4^ as well as MRI acquisition and post-processing (cortical reconstruction and lesion segmentation) have been described previously.^5,6^ Ethics approval was obtained from the Norwegian Regional Committees for Medical and Health Research Ethics (REK-814351, REK-2016/1906). All participants gave their written informed consent. All methods were performed in accordance with the relevant guidelines and regulations (Declaration of Helsinki).

For replication analyses, we used mixed and cross-sectional linear models to predict continuous variables (EDSS, PASAT, T2 lesions, brain volume) from log-transformed ALA. We used a negative binomial model to predict the occurrence of new MRI-T1 lesions and relapses. Both analyses were executed with and without the covariates mentioned in the original article^1^ (age, sex, clinical trial treatment arm) and for brain-derivatives additionally the estimated intracranial volume as a covariate to prevent distorting effects of head and brain size. Cross-sectional analyses of baseline data did not include treatment as a covariate, as no treatment had yet been administered in the historical clinical trial used.

For additional longitudinal estimations, similar to the original article, we examined the predictive effect of baseline ALA on the 12-year annual rate of change in EDSS, PASAT, T2 lesions number, brain volume, and the total number of T1w lesions and relapses at the 12-year follow-up. For additional exploratory analyses, we estimated the mediating effect of ALA for the relationship between a) brain volume and EDSS, and b) number of T2 lesions and EDSS using causal mediation analysis.^7^ Finally, as an additional quality control step, we used the disease duration as a covariate replacing age in the aforementioned analyses. All results are presented as standardized regression coefficients with 95% confidence intervals. We used an unadjusted alpha-level of 0.05.

## Results

Descriptive statistics across assessment time points can be found in Table 1. ALA measures presented high reliability, indicated by an intra-class correlation of κ=0.83 [0.77, 0.88], p < 0.00001.

**Table 1.** Baseline and 24-months follow-up characteristics of the cohort. The first 24 months of the study represent the period where ALA samples were collected.

|  | Baseline | 12-month follow-up | 24-month follow-up | 144-months follow-up |
| --- | --- | --- | --- | --- |
| Age | 38.8±8.3 | 39.8±8.3 | 40.8±8.3 | 49.6±8.46 |
| Females | 54 | 54 | 54 | 54 |
| Males | 31 | 31 | 31 | 31 |
| New relapses | 44 | 21 | 0 | 46 |
| Disease duration | 1.7±2.9 | 2.9±3.0 | 3.9±3.1 | 13.7±3.3 |
| ALA, $\mu\text{mol/dL}$ | 2.0±0.8 | 1.8±0.8 | 1.8±0.7 | - |
| # new T1 lesions | 0.5±0.5 | 0.1±0.3 | 0.2±0.4 | 0.5±0.5 |
| Total T2 lesions | 12.6±10.2 | 12.8±10.1 | 13.4±9.7 | 12.6±10.1 |
| EDSS | 1.9±0.8 | 2.1±1.1 | 2.2±1.3 | 2.5±1.3 |
| PASAT | 47.4±9.7 | 51.9±8.3 | 51.7±8.4 | 47.4±9.7 |

In cross-sectional data, looking at each time point individually, when controlling for age, sex, and treatment arm (omega-3 intake vs placebo), higher ALA levels were associated with lower EDSS at follow-up at month 24 (β = −0.68 [−1.22, −0.14], p = 0.013), and with the number of T2 lesions at baseline (β = 0.77 [0.14, 1.40], p = 0.017). ALA was also univariately, associated with lower EDSS at 12-months (β = −0.64 [−1.27, −0.01], p = 0.045) and 24-months follow-ups (β = −0.69 [−1.23, −0.14], p = 0.013). These associations were robust to exchanging the covariates age with disease duration except for the association between T2 lesions and ALA (p=0.104).

In longitudinal data over 24 months, multivariable mixed linear models adjusting for age, sex, treatment arm, and intra-cranial volume, higher ALA was significantly associated with greater brain volume, and lower disability, as measured with the EDSS-score (Table 2). Unexpectedly, a positive association was found between ALA and the number of T2 lesions.

**Table 2.** Standardized adjusted and unadjusted regression coefficients of log-transformed ALA. All multivariable models include the covariates age, sex, treatment arm, in addition to estimated intra-cranial volume when predicting T2 lesions and brain volumes. For quality control, in an additional set of analyses, age was replaced by disease duration. *This effect was non-significant (p=0.08) when including disease duration as a time-varying covariate instead of age.

| Outcome variable | Standardized Regression Coefficient<br>[95% Confidence Interval] | p-value | Model type |
| --- | --- | --- | --- |
| Brain volume | 0.26 [0.11, 0.41] | 0.0007 | Univariate |
|  | 0.22 [0.09, 0.36] | 0.00004 | Multivariable |
| # T2 lesions | 0.10 [-0.09, 0.28] | 0.31 | Univariate |
|  | 0.31 [0.03, 0.60] | 0.034* | Multivariable |
| EDSS | -0.37 [-0.63, -0.12] | 0.0037 | Univariate |
|  | -0.41 [-0.73, -0.08] | 0.012 | Multivariable |

Explorative cross-sectional causal mediation analyses at each time point did not suggest ALA as a mediator of the relationship between total brain volume and EDSS, or the number of T2 lesions and EDSS, as indicated by a non-significant proportion of mediated effects (p>0.05).

Baseline ALA was not associated with the annual rate of change between baseline and 12-year follow-up in PASAT, EDSS, number of T2 lesions, brain volume, relapses, or the total number of T1w lesions at the 12-year follow-up (p>0.05). The only notable, yet non-significant relationship was found for the annual relapses between baseline and 12-year follow-up (uncorrected β = −0.48 [−1.06, 0.96], p = 0.108; corrected β = −0.52 [−1.11, 0.08], p = 0.088).

## Discussion

In this replication study, we examined the relationship between serum ALA and long-term clinical and radiological outcomes in MS. Consistent with the findings by Cortese and collegues,^1^ we observed that higher levels of ALA were associated with lower disability and greater brain volume over the first two years of follow-up. These associations remained robust across univariate and multivariable models and demonstrated strong intra-individual stability over time. This suggest that higher ALA may be linked to more favourable disease profiles in MS, reinforcing its potential value as a biomarker. However, as highlighted in the original study,^1^ ALA lacked predictive power of long-term effects.

Our findings replicate the reported associations between ALA and lower disability as well as preserved brain structure. The consistency across independent cohorts strengthens the notion that ALA may play a protective role in MS-related neurodegeneration. However, several discrepancies also emerged. We found a positive association between ALA and T2-weighted lesion counts at baseline in multivariable models. This was not observed in the original study^1^ and contrasts previous findings from an earlier analysis of the same dataset,^6^ where higher ALA was associated with *fewer* new T2 lesions. Note, however that in the earlier analysis^6^ T2 lesions were manually evaluated, whereas the lesion counts presented here were automatically segmented. The instability of this result to disease duration as a covariate suggests a type I error. Our exploratory mediation analyses further indicated that ALA did not mediate the relationship between structural brain pathology and disability, suggesting that ALA may influence MS outcomes via independent pathways rather than acting as a mechanistic mediator.

A key strength of our study lies in the availability of independent, longitudinal clinical trial data^2^ with a low attrition rate.^5^ The repeated, highly reliable ALA measurements over two years, together with extended 12-year clinical and MRI follow-up, provided an opportunity to test both short- and long-term associations. However, our study also has important limitations. The sample size was modest, and not all participants had complete data at all time points, reducing statistical power for detecting small effects. Similar to the original study,^1^ we lacked follow-up serum ALA measurements beyond the initial two years, limiting our ability to model cumulative exposure or time-varying effects. Parts of the dataset, including MRI and dietary biomarker data, have been used in earlier publications^2,5,6^ of different analysts applying different methods. While this limits novelty, it offers a crucial validation step, as analyst choices influence MS research results.^8^

Taken together, our results support and extend prior findings linking serum ALA to reduced disability and preserved brain volume in MS. These findings add to a growing body of evidence pointing to the potential neuroprotective role of plant-based omega-3 fatty acids. Given the known anti-inflammatory^9,10^, membrane-stabilizing, and possible anti-viral properties of ALA,^11^ it is plausible that such mechanisms contribute to reduced MS activity, for example by interacting with immune processes related to viruses such as Epstein-Barr, which has recently been identified as a central etiological factor in MS.^12^ Despite these promising results, the lack of predictive power of ALA on long-term outcomes call for cautious interpretation. Larger, multicentre studies with repeated ALA sampling over extended time frames are needed to clarify causal pathways and establish the prognostic utility of ALA in MS. Future work should also explore whether ALA-related benefits generalize to other neuroinflammatory or neurodegenerative diseases, as suggested by recent evidence in amyotrophic lateral sclerosis (ALS).^13,14^

## Data availability

Data access can be requested after obtaining ethics approval. Analysis code is available on GitHub: https://github.com/MaxKorbmacher/ALA.

